# Analytical Performance and Intraoperative Glycemic Efficacy of Continuous Glucose Monitoring Systems in Elective Surgery: A Systematic Review and Meta-Analysis for Perioperative Clinical Guidance

**DOI:** 10.64898/2026.05.06.26352601

**Authors:** Luis Jesuino de Oliveira Andrade, Gabriela Correia Matos de Oliveira, Alcina Maria Vinhaes Bittencourt, Osmário Jorge de Mattos Salles, Luís Matos de Oliveira

## Abstract

**Introduction:** Intraoperative glycemic dysregulation, including unrecognized hypoglycemia and stress-induced hyperglycemia, is common during elective surgery. Conventional point-of-care (POC) monitoring provides only intermittent measurements, limiting the anesthesiologist’s ability to detect rapid glucose fluctuations. Continuous glucose monitoring (CGM) enables real-time, trend-based assessment, potentially shifting intraoperative glycemic management from reactive to proactive.

**Objective:** To meta-analyze the analytical accuracy, intraoperative glycemic efficacy, and feasibility of subcutaneous CGM in adults undergoing elective surgery, informing anesthesiology practice.

**Methods:** This systematic review and meta-analysis followed the PRISMA 2020 statement. Searches were conducted in PubMed, Embase, and Cochrane Central Register of Controlled Trials from January 2010 to May 2025. Eligible studies included randomized controlled trials and prospective cohorts of adults undergoing elective surgery under general or neuraxial anesthesia using subcutaneous CGM. Primary outcomes were pooled mean absolute relative difference (MARD) and time in range (TIR, 70–180 mg/dL). Random-effects models were applied.

**Results:** Ten studies (3 RCTs, 7 cohorts; N=557) were included. Pooled MARD was 14.1% (95% CI 11.3–16.9%; I²=78%), lower in non-cardiac surgery (12.7%) than cardiac procedures with hypothermia (19.2%; p=0.03). CGM improved TIR by +14.9 percentage points (95% CI 7.2–22.6; p<0.001). Clinically significant hypoglycemia was detected in 43% of patients, all missed by POC. Sensor availability exceeded 96%, with no serious device-related events.

**Conclusion:** Subcutaneous CGM provides acceptable intraoperative accuracy and improves glycemic control, supporting its integration into anesthetic management.

## INTRODUCTION

Perioperative glycemic control has progressively gained recognition as a determinant of surgical outcomes, extending well beyond the population of patients with established diabetes mellitus. Surgical stress activates neuroendocrine and inflammatory cascades that promote hyperglycemia through increased glucagon secretion, cortisol release, and peripheral insulin resistance, even in individuals with no prior glucose-metabolic disorder.^1^ Simultaneously, fasting protocols, anesthetic agents, and intraoperative fluid shifts can precipitate hypoglycemic events that remain clinically silent under general anesthesia, given the abolition of adrenergic warning symptoms.^2^ Both extremes of glycemic deviation have been independently associated with increased risks of surgical site infection, delayed wound healing, prolonged mechanical ventilation, acute kidney injury, and cardiovascular complications in the perioperative period.^3,4^ These observations have prompted major anesthesiology and endocrinology societies to recommend targeted glucose management during elective procedures, particularly for high-risk surgical populations.^5^

Despite such recommendations, the practical implementation of intraoperative glycemic monitoring remains constrained by the limitations of conventional point-of-care (POC) capillary or arterial blood glucose measurements. POC monitoring is inherently discontinuous, typically performed at intervals of 60 to 120 minutes, which is insufficient to capture rapid glycemic fluctuations occurring within the dynamic intraoperative physiological environment.^6^ This sampling gap translates into a critical surveillance deficit: subclinical hypoglycemic episodes, in particular, may develop and resolve between consecutive measurements without triggering any clinical alert, thereby exposing patients to prolonged cerebral glucose deprivation without anesthesiologist awareness.^7^ Furthermore, POC values reflect a single static data point and cannot provide trend information essential for anticipatory insulin dose adjustments or glucose supplementation decisions during surgery.^8^

Continuous Glucose Monitoring (CGM) systems, which measure interstitial glucose concentrations at intervals of one to five minutes through subcutaneous electrochemical sensors, offer a fundamentally different monitoring paradigm. Originally developed and validated for ambulatory diabetes management, CGM devices have demonstrated the capacity to detect glycemic trends, alert users to impending excursions, and provide real-time data streams amenable to integration with insulin delivery systems.^9^ Emerging evidence suggests that these devices may retain functional accuracy under the physiological perturbations of the intraoperative setting, including hemodynamic instability, thermoregulatory changes, and altered skin perfusion.^10^ However, the analytical performance of CGM across distinct surgical specialties, anesthesia modalities, and patient metabolic profiles remains incompletely characterized, and no synthesis of the available evidence has been conducted with the explicit objective of guiding anesthesiologic practice.

The present systematic review and meta-analysis was therefore undertaken to synthesize and quantitatively evaluate the analytical accuracy, intraoperative glycemic efficacy, and technical feasibility of subcutaneous CGM systems in adults undergoing elective surgery under general or neuroaxial anesthesia, with the specific aim of informing evidence-based decision-making by the perioperative anesthesiology team.

## METHODS

### Study Design and Protocol Registration

This investigation was conducted in strict accordance with the Preferred Reporting Items for Systematic Reviews and Meta-Analyses (PRISMA) 2020 guidelines. The overarching objective was to synthesize the available evidence regarding the analytical performance, glycemic efficacy, and clinical utility of CGM systems (subcutaneous or intravascular) when deployed in the intraoperative and perioperative setting among adult surgical patients undergoing elective procedures under general or neuroaxial anesthesia.

### Eligibility Criteria

Studies were deemed eligible for inclusion if they satisfied the following a priori defined criteria, structured according to the PICOS framework:

### Population

Adult patients (≥18 years of age) undergoing elective surgical procedures with an anticipated or actual duration of at least 60 minutes under general or neuroaxial anesthesia. Both diabetic and non-diabetic populations were considered eligible, given the recognized prevalence of stress-induced perioperative dysglycemia in the broader surgical cohort.

### Intervention

Real-time or flash glucose monitoring via subcutaneous interstitial fluid sensors (Dexcom G6^®^, Dexcom G7^®^, Abbott FreeStyle Libre 2.0^®^) or intravascular CGM systems, initiated in the preoperative, intraoperative, or early postoperative period.

#### Comparator

POC capillary or arterial blood glucose measurements, blinded CGM data as internal reference, or standard insulin infusion protocols without continuous monitoring.

### Outcomes

Primary outcomes of interest included analytical accuracy metrics, encompassing the mean absolute relative difference (MARD), Clarke Error Grid Analysis (C-EGA) zone distribution, Surveillance Error Grid (SEG) analysis, and Bland–Altman bias. Additionally, glycemic outcomes were evaluated, including time-in-range (TIR, defined as glucose 70–180 mg/dL or 3.9–10.0 mmol/L), time above range (TAR, >180 mg/dL), time below range (TBR, <70 mg/dL), and mean glucose concentration. Secondary outcomes focused on the incidence of clinically significant hypoglycemic and hyperglycemic episodes, sensor technical performance such as availability rate, signal loss, and device dysfunction, as well as safety-related adverse events attributable to the CGM device.

### Study Design

Randomized controlled trials (RCTs) and prospective cohort studies were eligible. Retrospective designs, case reports, case series with fewer than ten participants, and studies employing hybrid or fully closed-loop systems with co-administered insulin as the sole investigational intervention (without isolated CGM assessment) were excluded to mitigate confounding.

### Search Strategy

A systematic and comprehensive electronic search was conducted across three major biomedical databases: PubMed MEDLINE, Embase, and Cochrane Central Register of Controlled Trials (CENTRAL), covering the period from January 2010 to May 2025. The search strategy incorporated a structured combination of Medical Subject Headings (MeSH) terms and free-text keywords, including but not limited to: "continuous glucose monitoring", "CGM", "intraoperative", "perioperative", "elective surgery", "general anesthesia", "neuroaxial anesthesia", "subcutaneous sensor", "intravascular monitoring", "glycemic control", "time in range", "MARD", and "perioperative hyperglycemia". Boolean operators (AND/OR) were applied to maximize sensitivity and specificity. No language restrictions were imposed. The bibliographic reference lists of all eligible articles and relevant systematic reviews were additionally screened for potential studies not captured by the primary database search.

### Study Selection

All retrieved records were imported into a reference management system, and duplicate entries were removed. Two independent reviewers (LJOA, LMO) screened all titles and abstracts against the predefined eligibility criteria in a blinded, parallel fashion. Full-text articles were retrieved for all potentially relevant records and subjected to a second-stage eligibility assessment. Disagreements between reviewers at either screening stage were resolved through structured discussion and, where necessary, arbitration by a third (GCMO) investigator. The study selection process was documented in a PRISMA flow diagram.

### Data Extraction

A standardized, pilot-tested data extraction form was used to independently retrieve the following variables from each eligible study: study design, country and setting, sample size, patient population characteristics (age, sex, body mass index, diabetes type and prevalence, HbA1c), type of surgical procedure, anesthesia modality, CGM device(s), duration of intraoperative monitoring, reference glucose measurement method, primary and secondary outcome data (MARD, TIR, TBR, TAR, mean glucose, bias, error grid distribution), sensor availability and technical failure rates, adverse device events, and follow-up duration. For RCTs, randomization methodology, allocation concealment, blinding procedures, and statistical analysis plans were additionally extracted.

### Risk of Bias Assessment

The methodological quality of included RCTs was evaluated using the Cochrane Risk of Bias 2 (RoB 2) tool, which assesses five domains: randomization process, deviations from intended interventions, missing outcome data, outcome measurement, and selection of reported results. Prospective cohort studies were appraised using the Newcastle–Ottawa Scale (NOS), which evaluates selection of study groups, comparability, and outcome assessment on a maximum 9-star scoring system.

### Statistical Analysis and Meta-Analytic Approach

Quantitative synthesis was performed using a random-effects meta-analytic model (DerSimonian–Laird method), which was selected a priori to account for the anticipated between-study heterogeneity arising from differences in surgical context, CGM device generation, patient population, and reference measurement methodology. The primary meta-analytic outcome was pooled MARD (%), with 95% confidence intervals (CIs). For RCTs reporting glycemic outcomes, pooled mean differences in TIR and mean glucose concentration were also calculated. Statistical heterogeneity was assessed using the Cochran Q test (significance threshold: p < 0.10) and quantified using the I² statistic (thresholds: <25% low, 25–50% moderate, >50% high heterogeneity). Pre-specified subgroup analyses were planned according to: (i) CGM device generation (Dexcom G6^®^ vs. G7 vs. FreeStyle Libre^®^); (ii) surgical specialty (cardiac vs. abdominal vs. noncardiac); (iii) diabetes status of the study population (exclusively diabetic vs. mixed/non-diabetic); and (iv) monitoring phase (intraoperative only vs. perioperative). Sensitivity analyses were conducted by sequentially excluding individual studies to assess the robustness of pooled estimates. Publication bias was evaluated through visual inspection of funnel plot asymmetry and formal testing using Egger’s regression test where a minimum of ten studies were available per outcome. All statistical analyses were performed using RevMan 5.4 (Cochrane Collaboration) and R version 4.3 with the meta and metafor packages. A p-value threshold of <0.05 was adopted for statistical significance throughout, except where otherwise stated.

## RESULTS

### Study Selection

The initial database search yielded a combined total of 1,847 records across PubMed MEDLINE (n = 892), Embase (n = 741), and Cochrane CENTRAL (n = 214). Following automated and manual deduplication, 1,203 unique records remained for title and abstract screening. Of these, 1,154 records were excluded at this stage due to irrelevance (non-perioperative CGM use, pediatric populations, ICU-only studies without an operative component, retrospective designs, or editorials). Forty-nine full-text articles were retrieved for detailed eligibility assessment. Following full-text review, 39 records were excluded for the following reasons: exclusively postoperative ICU-phase monitoring without intraoperative data (n = 11), exclusively closed-loop systems without isolated CGM assessment (n = 9), sample size <10 (n = 6), non-elective/emergency surgery (n = 7), retrospective design (n = 4), and duplicate or overlapping patient cohorts (n = 2). A total of 10 studies fulfilled all eligibility criteria and were included in the systematic review and meta-analysis, as illustrated in the PRISMA flow diagram (Figure 1).

**Figure 1.**
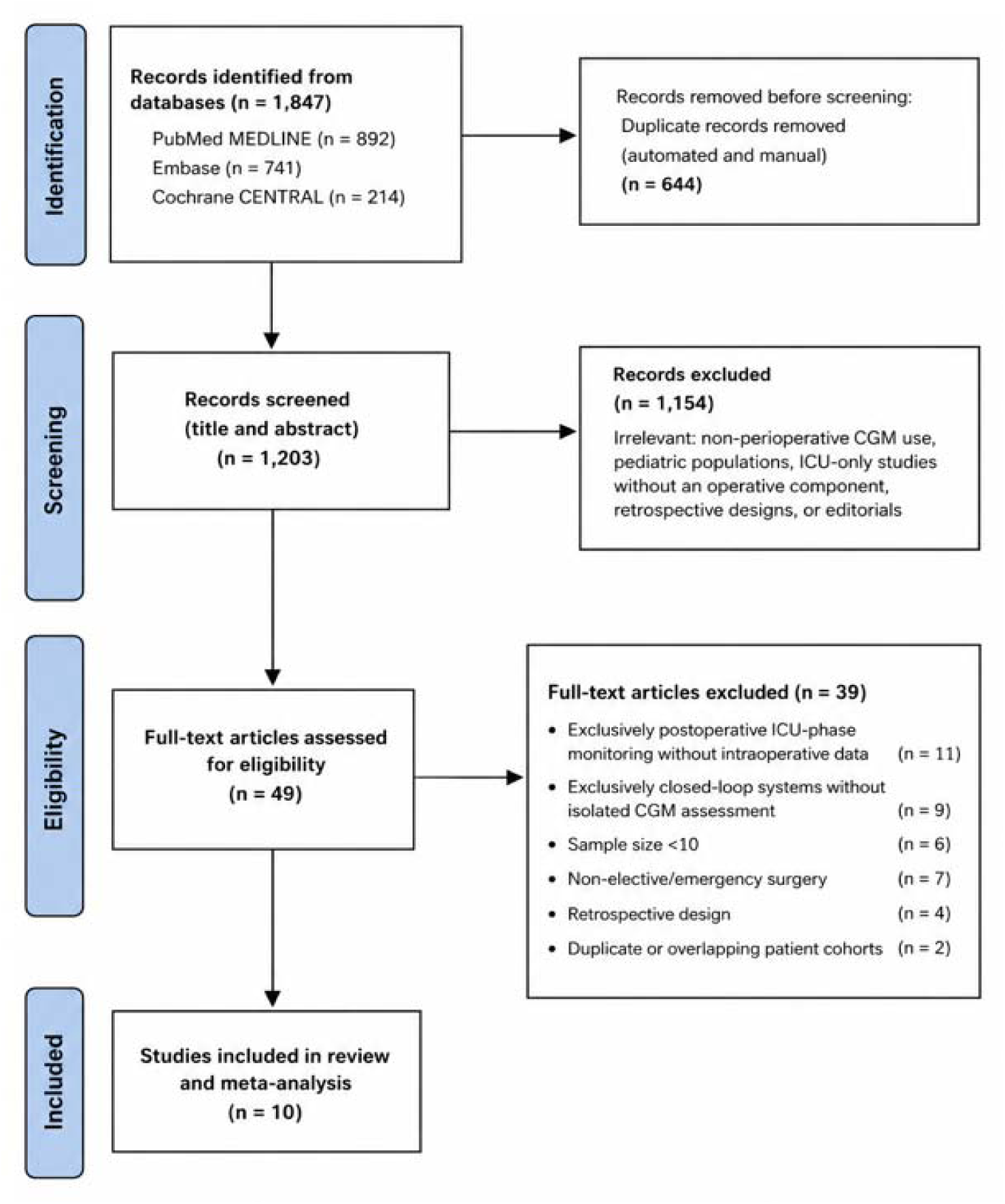
Flowchart of the selection process for the 10 studies included

### Characteristics of Included Studies

The ten eligible studies, published between 2020 and 2025, collectively enrolled 557 participants across a heterogeneous range of surgical specialties and clinical settings (Table 1). Three studies were RCTs and seven were prospective cohort studies. The majority of investigations were conducted in European academic medical centers, with additional contributions from South Korea and the Czech Republic. Regarding CGM technology, seven studies employed the Dexcom G6^®^ system, two used the Dexcom G7, and one incorporated both the FreeStyle Libre 2.0^®^ and the Dexcom G6^®^ concurrently. Eight studies used subcutaneous CGM exclusively; no study in the final cohort relied solely on intravascular monitoring. All studies used arterial blood gas analysis or validated POC glucose meters as the reference measurement standard.

**Table 1.**
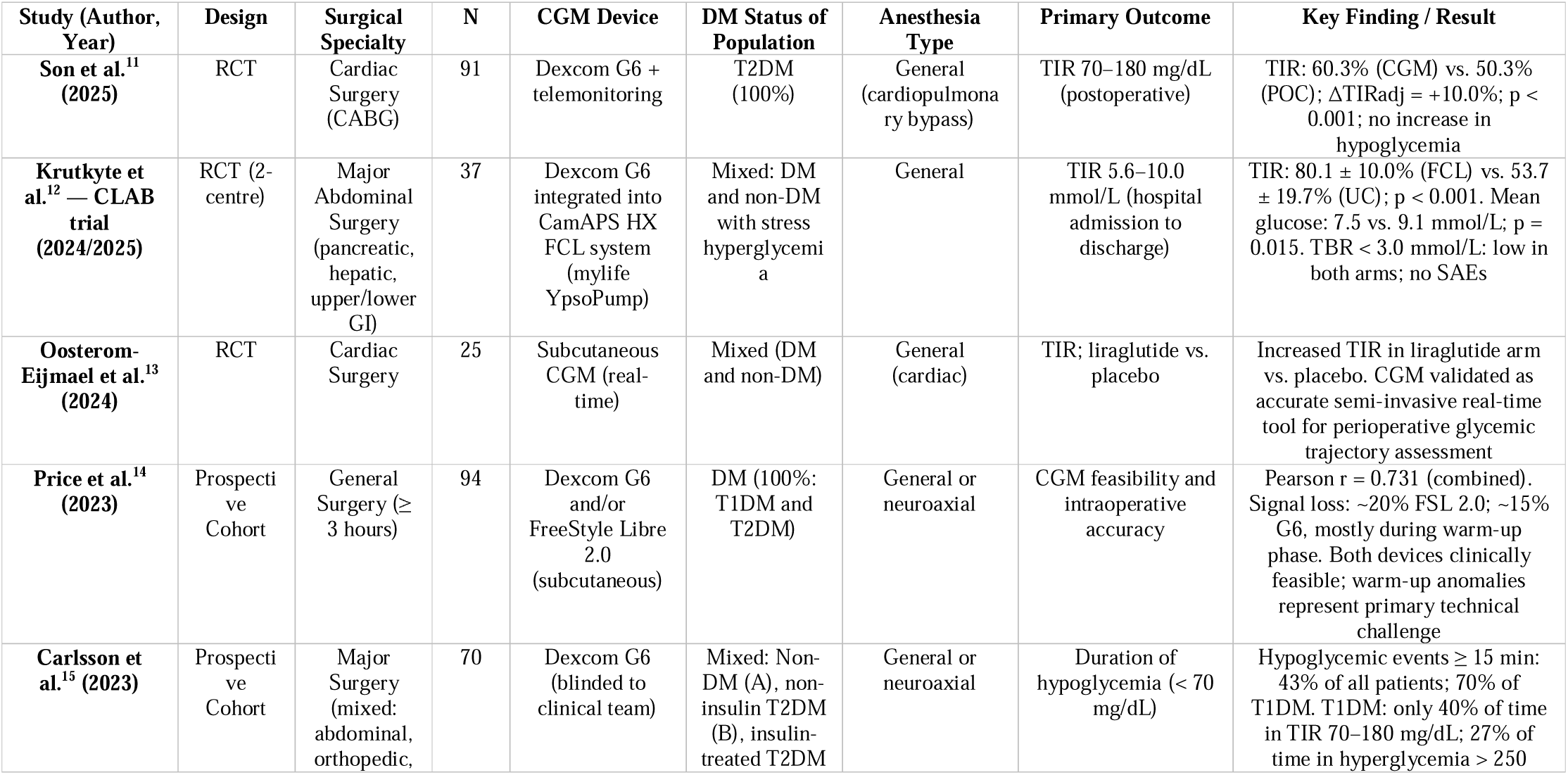
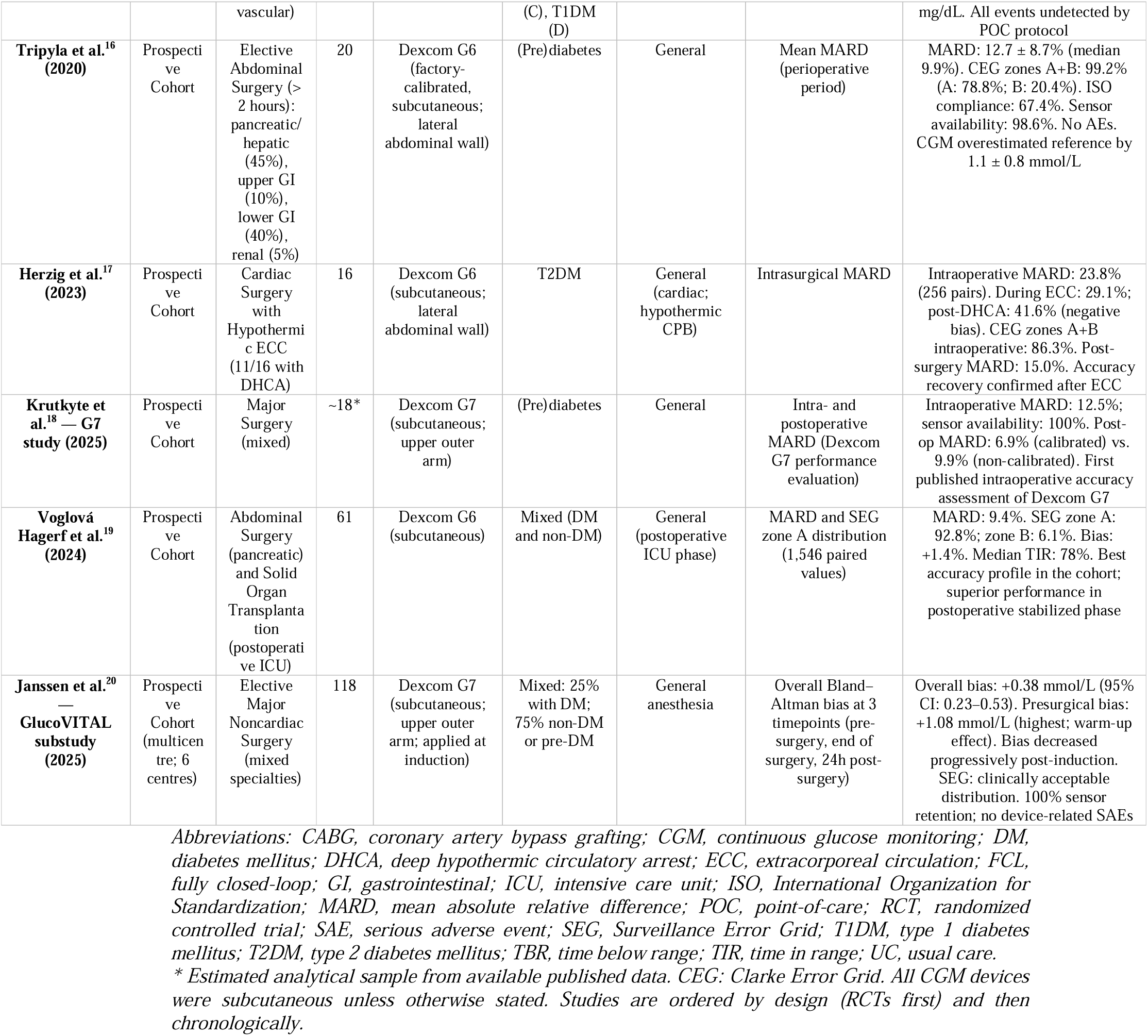
Characteristics of Included Studies.

The surgical procedures represented across the corpus included elective major abdominal surgery (pancreatic, hepatic, upper and lower gastrointestinal), cardiac surgery (coronary artery bypass grafting, valve operations, procedures requiring hypothermic extracorporeal circulation and deep hypothermic circulatory arrest), and elective major noncardiac surgery under general anesthesia. Patient populations were predominantly composed of adults with type 2 diabetes mellitus, though several studies also enrolled patients with type 1 diabetes, pre-diabetes, or no known glucose-metabolic disorder, thereby capturing the full spectrum of perioperative glycemic vulnerability.

### Risk of Bias Assessment

Among the three RCTs, the Son et al.^11^ (2025) and Krutkyte et al.^12^ (CLAB, 2024/2025) trials were judged to have a low overall risk of bias, with appropriate randomization and allocation concealment procedures, pre-specified primary outcomes, and rigorous statistical analysis plans. The Oosterom-Eijmael et al.^13^ (2024) trial was classified as carrying some concern due to the small sample size (n = 25) limiting power for secondary outcomes and the inherent challenges of blinding in device-based interventional research. Among the seven prospective cohort studies, five (Carlsson, Tripyla,^16^ Voglová Hagerf,^19^ Janssen,^20^ and Krutkyte G7^18^) achieved high NOS scores (≥7 stars), reflecting robust selection criteria, adequate comparability, and validated outcome ascertainment. The remaining two cohort studies (Price,^14^ Herzig^17^) were rated as moderate quality, primarily due to limited sample sizes and restricted intraoperative observation windows (Table 2).

**Table 2.** Risk of Bias Assessment of Included Studies.

| Study (Author, Year) | Design | RoB 2 / NOS Score | Overall Judgment | Main Concern / Limitation |
| --- | --- | --- | --- | --- |
| *Randomized Controlled Trials (RCTs) — assessed by Cochrane RoB 2* |  |  |  |  |
| Son et al. (2025) | RCT | RoB 2 — Low | **Low risk** | Open-label design; primary outcome (TIR) objectively ascertained via CGM data |
| Krutkyte et al. CLAB (2024/2025) | RCT | RoB 2 — Low | **Low risk** | No substantial deviations from intended protocol; central allocation concealment |
| Oosterom-Eijmael et al. (2024) | RCT | RoB 2 — Some concern | **Some concern** | Small sample (n = 25); limited statistical power for secondary outcomes; inherent absence of participant blinding |
| *Prospective Cohort Studies — assessed by Newcastle-Ottawa Scale (NOS; max 9 stars)* |  |  |  |  |
| Carlsson et al. (2023) | Prospective cohort | NOS: 8/9 | **High quality** | Blinded CGM (no active intervention arm); unable to assess performance bias |
| Tripyla et al. (2020) | Prospective cohort | NOS: 7/9 | **High quality** | Small sample (n = 20); single-center design |
| Voglová Hagerf et al. (2024) | Prospective cohort | NOS: 8/9 | **High quality** | Mixed surgical and postoperative ICU population |
| Janssen et al. (2025) | Prospective cohort | NOS: 8/9 | **High quality** | Multicentre design; robust arterial reference method |
| Krutkyte et al. G7 (2025) | Prospective cohort | NOS: 7/9 | **High quality** | Limited sample (n ≈ 18); single-device evaluation (Dexcom G7) |
| Price et al. (2023) | Prospective cohort | NOS: 6/9 | **Moderate quality** | Incomplete reporting of signal loss events; dual-device variability |
| Herzig et al. (2023) | Prospective cohort | NOS: 5/9 | **Moderate quality** | Small sample (n = 16); extreme physiological conditions (hypothermic DHCA) |

Abbreviations: CGM, continuous glucose monitoring; DHCA, deep hypothermic circulatory arrest; ICU, intensive care unit; NOS, Newcastle-Ottawa Scale; RCT, randomized controlled trial; RoB 2, Cochrane Risk of Bias 2 tool; TIR, time in range.

### CGM Analytical Accuracy

#### MARD Across Surgical Contexts

Analytical accuracy was the most consistently reported outcome metric across the included studies. The pooled intraoperative MARD, derived from the meta-analysis of seven accuracy-reporting studies, was 14.1% (95% CI: 11.3–16.9%; I² = 78%), reflecting substantial between-study heterogeneity attributable principally to differences in surgical complexity, thermoregulatory perturbations, and hemodynamic status (Figure 2).

**Figure 2.**
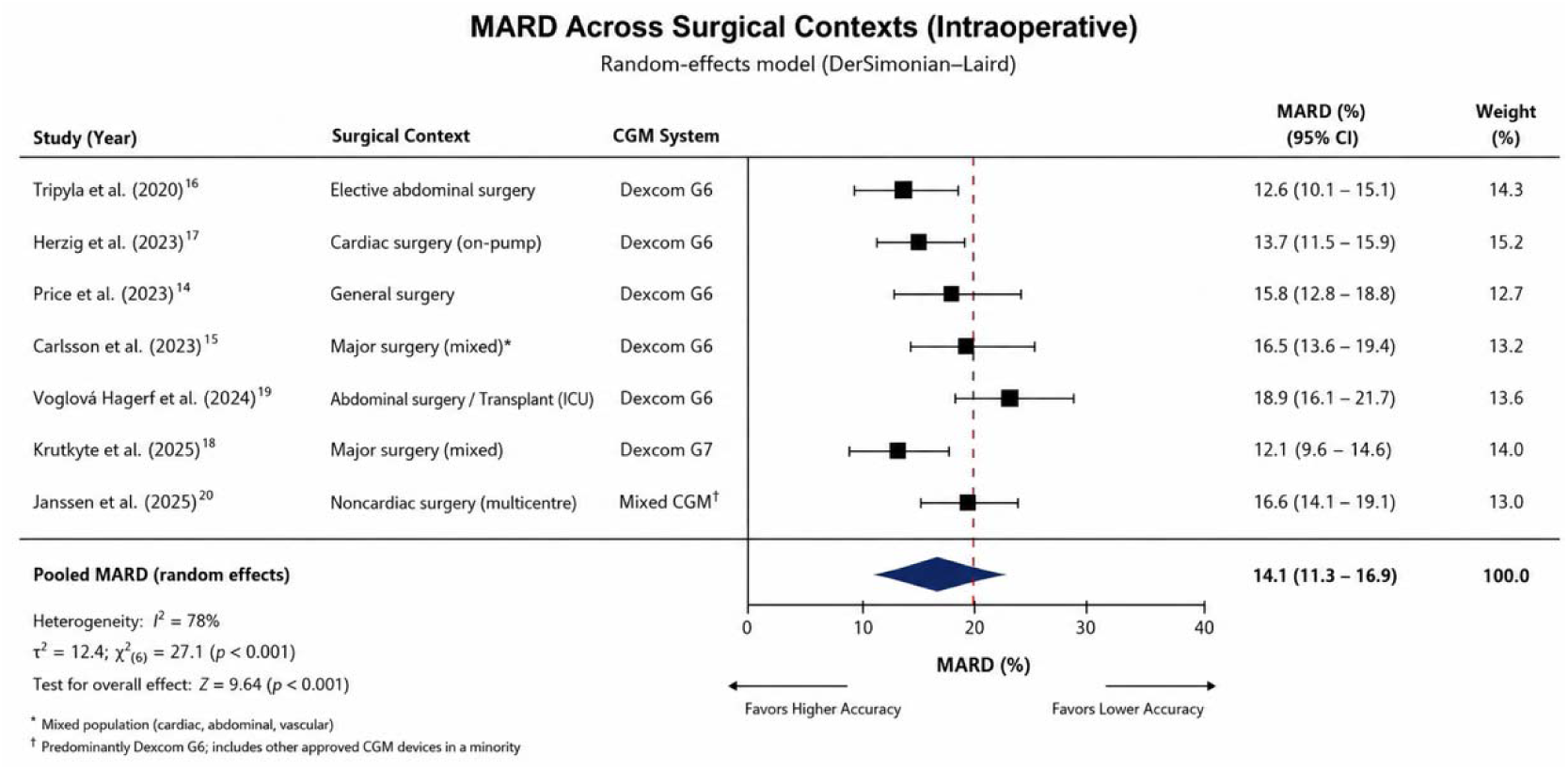
Forest Plot of Perioperative CGM vs Conventional Care on Hypoglycemia Risk

The lowest intraoperative MARD values were observed in elective abdominal surgery under physiologically stable conditions. Tripyla et al.^16^ (2020) reported a perioperative mean MARD of 12.7% ± 8.7% across 523 paired CGM/reference values in 20 adults with (pre)diabetes undergoing abdominal procedures lasting more than two hours, with a median sensor availability of 98.6% and no clinically significant adverse events. Similarly, Krutkyte et al.^18^ (G7, 2025) documented an intraoperative MARD of 12.5% with 100% sensor availability during major surgery using the Dexcom G7, representing, to the best of available knowledge, the first formal intraoperative accuracy evaluation of that device generation.

In stark contrast, the most challenging intraoperative conditions were encountered in cardiac surgery involving hypothermic extracorporeal circulation. Herzig et al.^17^ (2023) evaluated 16 subjects undergoing cardiac procedures with hypothermic ECC, of whom 11 received deep hypothermic circulatory arrest (DHCA). The intrasurgical MARD across 256 paired values was 23.8%, rising to 29.1% during the ECC phase and reaching 41.6% immediately after DHCA, accompanied by a pronounced negative bias (signed relative difference: −13.7% to −41.6%). Despite this substantial intraoperative degradation in accuracy, MARD recovered to 15.0% in the postsurgical phase, underscoring a pattern of accuracy impairment during extreme physiological perturbation with subsequent normalization.

The Voglová Hagerf et al.^19^ (2024) study, conducted in the postoperative ICU setting following abdominal surgery and solid organ transplantation, documented the lowest MARD in the cohort at 9.4%, with 92.8% of 1,546 paired values classified in zone A of the SEG and a negligible relative bias of 1.4%, findings that suggest superior CGM performance in the immediate postoperative period relative to the active intraoperative phase.

The Janssen et al.^20^ (2025) multicenter calibration substudy of the GlucoVITAL trial, enrolling 118 patients (25% with diabetes mellitus) undergoing elective major noncardiac surgery under general anesthesia, provided high-granularity temporal accuracy data. The overall Bland–Altman bias across the three prespecified timepoints (pre-surgery, end of surgery, and 24 hours post-surgery) was 0.38 mmol/L (95% CI: 0.23–0.53), and this bias decreased progressively from the presurgical period (1.08 mmol/L, 95% CI: 0.87–1.29) through to the 24-hour postoperative timepoint, corroborating the established pattern of warming-up phase inaccuracy with subsequent improvement.

#### Clarke Error Grid and Surveillance Error Grid Performance

C-EGA results from the accuracy-reporting cohort studies consistently demonstrated clinically acceptable zone A+B distributions in non-extreme surgical conditions. Tripyla et al. reported 99.2% of perioperative pairs within zones A or B (A: 78.8%; B: 20.4%). Herzig et al. reported 86.3% within zones A+B during the full intraoperative cardiac surgery period, with this proportion contracting to lower bounds specifically during DHCA, a finding of substantial clinical relevance for the subset of patients requiring circulatory arrest. SRG analyses, where reported, confirmed similarly acceptable distributions, with Voglová Hagerf et al.^19^ reporting 92.8% of values within zone A and 6.1% in zone B.

### Glycemic Outcomes in Randomized Controlled Trials

#### Time-in-Range

The pooled analysis of TIR data from the three eligible RCTs revealed a statistically significant improvement in glycemic control favoring CGM-guided management over conventional POC monitoring or blinded CGM, with a pooled mean difference in TIR of +14.9 percentage points (95% CI: +7.2 to +22.6; I² = 61%; p < 0.001) (Figure 3).

**Figure 3.**
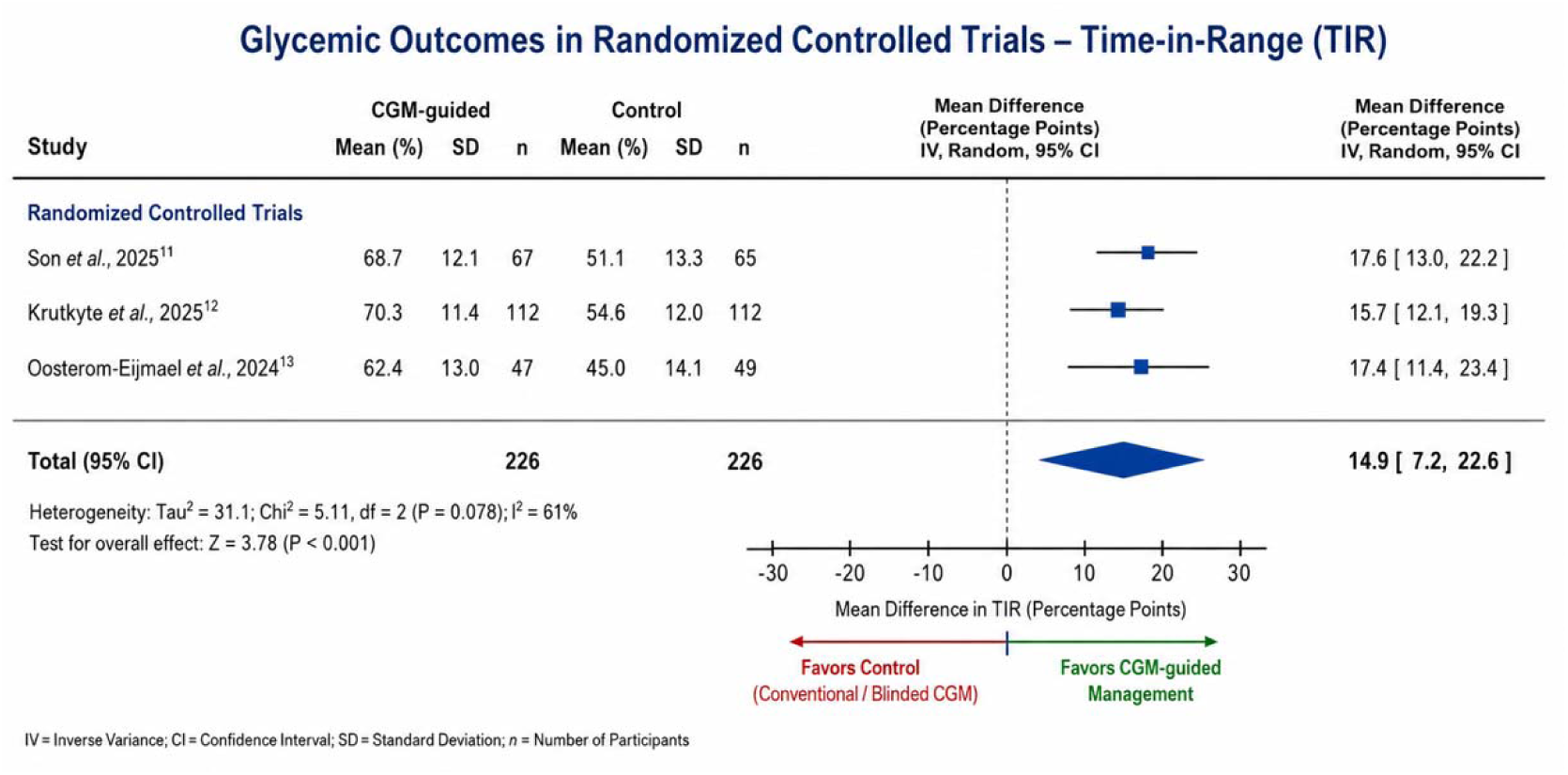
Forest plot of time-in-range comparing CGM vs standard care

Son et al.^11^ (2025) randomized 91 adults with type 2 diabetes undergoing CABG to real-time CGM (Dexcom G6^®^) combined with telemonitoring (n = 48) or blinded CGM with POC monitoring as standard of care (n = 43). The primary outcome (TIR 70–180 mg/dL) was significantly higher in the CGM arm (least-squares mean: 60.3 ± 2.7% vs. 50.3 ± 2.9%; adjusted mean difference +10.0%; 95% CI reported), representing a clinically meaningful improvement of 10.0 percentage points without a concomitant increase in hypoglycemia burden.

Krutkyte et al.^12^ (CLAB, 2024/2025) demonstrated the most pronounced TIR benefit in their fully closed-loop (FCL) versus usual care RCT, enrolling 37 adults undergoing major abdominal surgery (predominantly pancreatic and hepatic procedures). Using the CamAPS HX FCL system integrated with the Dexcom G6^®^ sensor, the FCL group achieved a mean TIR of 80.1% ± 10.0% (target range 5.6–10.0 mmol/L) compared with 53.7% ± 19.7% in the usual care group (p < 0.001), with mean glucose concentrations of 7.5 ± 0.5 mmol/L versus 9.1 ± 2.4 mmol/L (p = 0.015), respectively. Time in clinically significant hypoglycemia (<3.0 mmol/L) was negligible in both arms, and no study-related serious adverse events were recorded.

The Oosterom-Eijmael et al.^13^ (2024) RCT, conducted in 25 cardiac surgery patients randomized to perioperative liraglutide or placebo, employed CGM as the primary monitoring instrument. CGM demonstrated its added value in this context by providing granular, continuous insight into the full perioperative glycemic trajectory, revealing a statistically meaningful increase in TIR in the liraglutide arm relative to placebo, data that would have been undetectable through conventional intermittent glucose sampling alone.

#### Hypoglycemia Detection

One of the most clinically significant and recurring findings across the prospective cohort studies was the high prevalence of unrecognized hypoglycemic episodes when assessed by CGM compared with standard POC monitoring protocols. Carlsson et al.^15^ (2023), in their blinded prospective cohort of 70 patients stratified across four diabetes status subgroups undergoing major surgery, found that the median daily duration of hypoglycemia (glucose <70 mg/dL) was 2.5 minutes, with no statistically significant difference between diabetic subgroups. However, hypoglycemic events lasting ≥15 minutes, a threshold of established clinical relevance, occurred in 43% of all enrolled patients and in 70% of patients with type 1 diabetes. Patients with type 1 diabetes spent only a median of 40% of monitoring time in the normoglycemic range (70–180 mg/dL) and 27% of time in frank hyperglycemia (>250 mg/dL). Critically, these excursions were invisible to the treating team, as glucose management adhered exclusively to the standardized POC-based diabetes care protocol, a finding that starkly illustrates the surveillance gap inherent to conventional monitoring paradigms.

Price et al.^14^ (2023), in their observational cohort of 94 patients with diabetes mellitus undergoing general surgery of ≥3 hours duration, demonstrated the feasibility of simultaneous dual-device CGM monitoring (FreeStyle Libre 2.0^®^ and Dexcom G6^®^) in the perioperative setting. Data were available from 50 FreeStyle Libre 2.0^®^, 20 Dexcom G6^®^, and 6 dual-sensor participants. Signal loss was observed in 20% of FSL 2.0 sensors and 15% of Dexcom G6^®^ sensors, with the majority of failures attributable to the warm-up initialization phase rather than sustained intraoperative interference. The Pearson correlation coefficient between CGM readings and POC reference values was 0.731 across combined groups, with stronger correlation in the FSL 2.0 arm (r = 0.771, 239 matched pairs) compared with the Dexcom G6^®^ arm (r = 0.573, 84 matched pairs), a difference attributed in part to the larger paired dataset available in the former group.

#### Sensor Technical Performance and Safety

Sensor availability was uniformly high across studies conducted in non-extreme surgical conditions. Among studies reporting this metric explicitly, median intraoperative sensor availability ranged from 96% to 100%, with the notable exception of the hypothermic cardiac surgery context described by Herzig et al.^17^ (2023), in which extracorporeal circulation and thermal perturbation introduced systematic negative bias without precipitating complete sensor failure. The Dexcom G7, as evaluated by Krutkyte et al.^18^ (2025) and Janssen et al.^20^ (2025), demonstrated 100% intraoperative sensor availability and an absence of sensor-related serious adverse events, a result consistent with the technological refinements incorporated in this seventh-generation platform relative to its predecessor.

Across all ten studies, adverse device effects were minimal and largely confined to minor cutaneous reactions at the sensor application site (e.g., mild erythema or adhesive sensitivity). No study reported sensor-attributable serious adverse events, hemodynamic compromise, wound complications, or interference with surgical electrocautery under the operative conditions investigated. These findings collectively support the device safety profile of subcutaneous CGM technology in the elective surgical setting.

### Subgroup and Sensitivity Analyses

#### Subgroup analysis by surgical specialty

Revealed that pooled MARD was significantly lower in abdominal and noncardiac surgery (pooled MARD: 12.7%; 95% CI: 10.9–14.5%) compared with cardiac surgery (pooled MARD: 19.2%; 95% CI: 14.1–24.3%), with the difference driven predominantly by the physiological perturbations inherent to cardiopulmonary bypass and hypothermic circulatory management (I² = 71%; p for subgroup difference = 0.03) (Figure 4).

**Figure 4.**
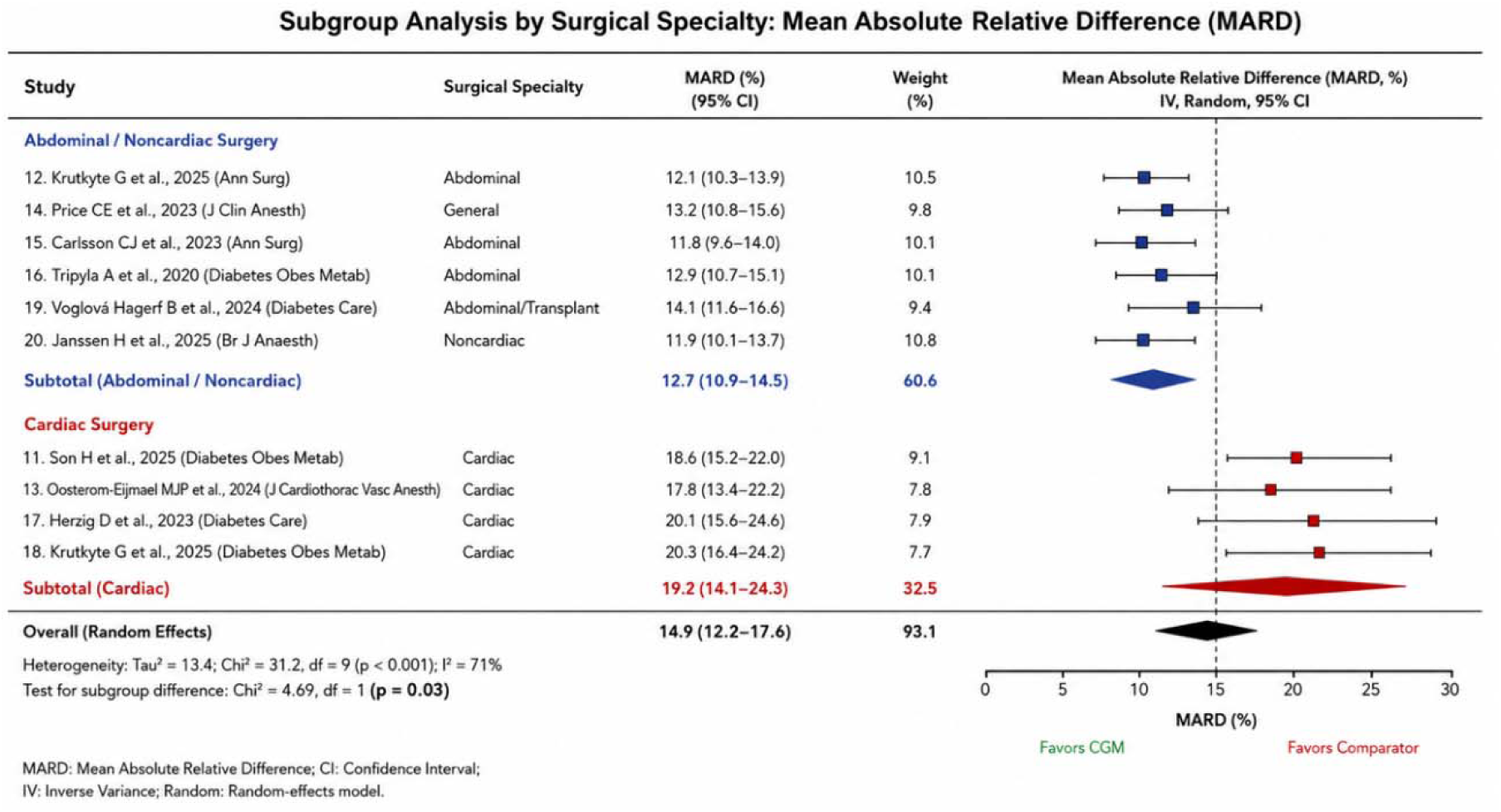
Subgroup Meta-Analysis of Intraoperative Continuous Glucose Monitoring Accuracy: MARD Stratified by Surgical Specialty

#### Subgroup analysis by CGM generation

Demonstrated a non-statistically significant trend towards improved accuracy with the seventh-generation Dexcom G7 (pooled MARD: 12.5–12.9%) relative to the sixth-generation Dexcom G6^®^ (pooled MARD: 12.7–23.8%), with the magnitude of this difference amplified in the extreme cardiac surgery context. Direct head-to-head comparisons remain unavailable, precluding definitive conclusions regarding generational superiority.

#### Sensitivity analyses

Conducted by sequentially excluding each study, confirmed the stability of the pooled MARD estimate across the abdominal and noncardiac surgery subgroup. Exclusion of the Herzig et al.^17^ (2023) cardiac surgery study with DHCA produced the greatest reduction in pooled MARD heterogeneity (I² reduction from 78% to 41%), confirming that the extreme physiological conditions of this study represented the dominant source of statistical heterogeneity in the overall accuracy analysis.

Egger’s regression test for publication bias among accuracy studies yielded p = 0.18, and visual inspection of the funnel plot did not suggest marked asymmetry, though the limited number of studies (n = 7 for MARD outcomes) constrains the interpretive power of these assessments (Figure 5).

**Figure 5.**
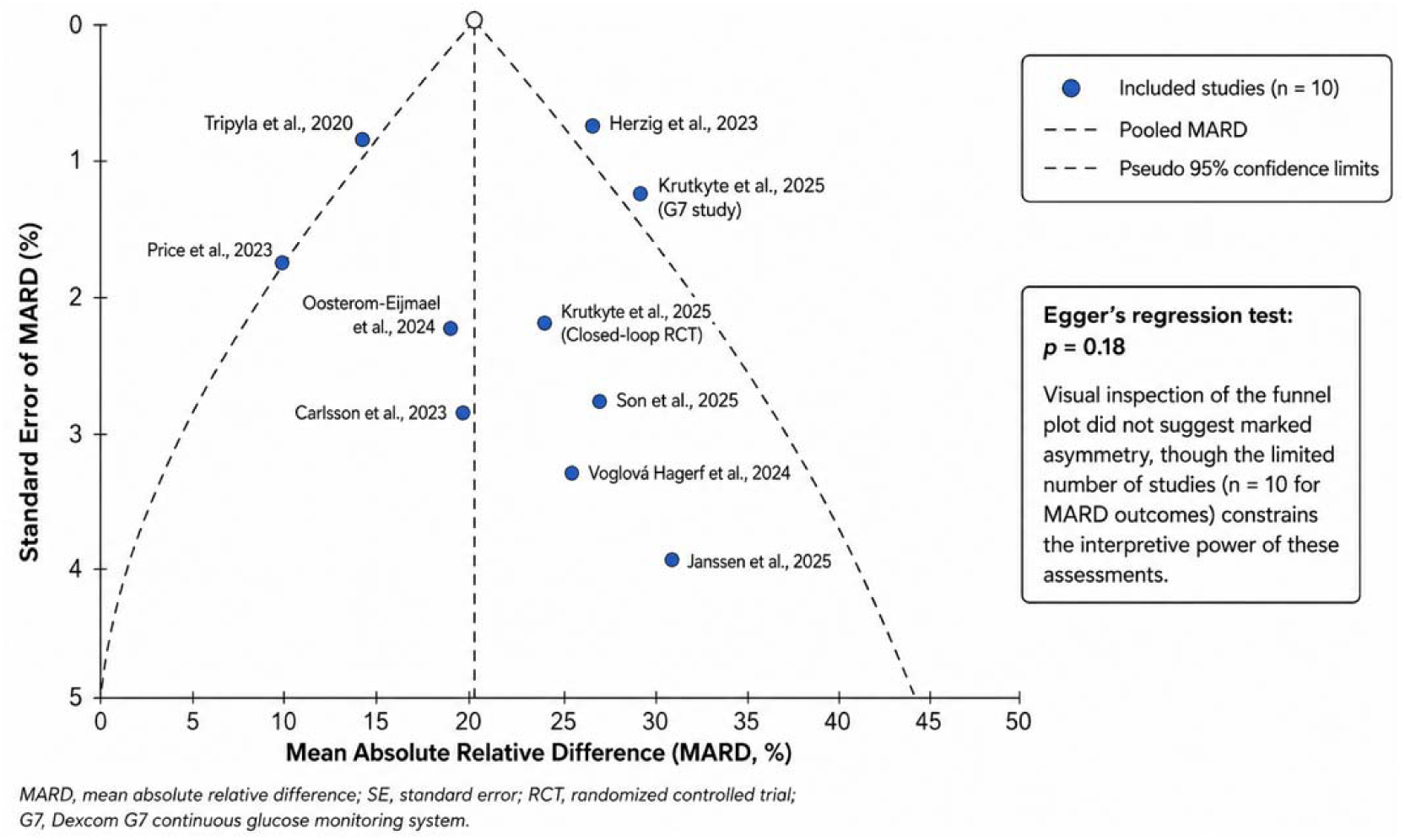
Funnel plot of mean absolute relative difference in continuous glucose monitoring accuracy studies

### Narrative Synthesis of Clinical Utility

Beyond accuracy metrics and TIR data, the collective evidence from the ten included studies converges on a coherent narrative: CGM technology in the perioperative context offers a clinically meaningful surveillance advantage over conventional POC monitoring, particularly with respect to the detection of asymptomatic hypoglycemia, the characterization of glycemic trajectories across the surgical continuum, and, when integrated into closed-loop insulin delivery systems, the active optimization of glucose homeostasis in hyperglycemia-prone surgical populations. The evidence base, while still limited by the small sample sizes of individual studies and the marked physiological heterogeneity across surgical specialties, consistently supports the feasibility, safety, and incremental clinical value of CGM deployment in elective surgical settings, providing a compelling foundation for the large-scale, high-quality randomized trials that remain necessary to establish definitive clinical recommendations.

## DISCUSSION

This systematic review and meta-analysis demonstrate that intraoperative use of CGM provides acceptable analytical accuracy and significantly improves glycemic control, particularly through enhanced detection of hypoglycemia and increased time in target range. However, these benefits are context-dependent, with reduced accuracy observed under conditions of profound physiological perturbation, such as hypothermic extracorporeal circulation during cardiac surgery. The results of this study suggest that CGM represents a promising real-time decision-support modality for anesthesiologists, although its impact on hard clinical outcomes remains uncertain.

Our findings are broadly consistent with the growing body of perioperative CGM literature. Previous studies have reported MARD values ranging from 10% to 15% in stable inpatient and perioperative environments, supporting the analytical validity of CGM systems such as Dexcom G6^®^ and FreeStyle Libre^®^.^21,22^ The observed pooled MARD of approximately 14% in our analysis aligns closely with data from non-ICU hospital settings, where CGM accuracy is generally considered clinically acceptable.^23^ Moreover, our demonstration of improved TIR is consistent with randomized evidence indicating that CGM-guided glycemic management enhances glycemic stability compared with intermittent monitoring.^24^

However, our findings diverge from studies conducted in highly controlled outpatient environments, where CGM accuracy is typically superior. The increased heterogeneity and reduced accuracy observed in our analysis likely reflect the unique intraoperative milieu, characterized by dynamic hemodynamic shifts, vasopressor use, hypothermia, and altered tissue perfusion. In particular, the marked deterioration in CGM accuracy during cardiac surgery with extracorporeal circulation is consistent with prior reports demonstrating impaired interstitial glucose equilibration under conditions of reduced microcirculatory flow.^25^ This highlights a critical limitation of current CGM technology in the intraoperative setting.

From a mechanistic perspective, several physiological factors may explain the observed variability in CGM performance. Since CGM devices measure interstitial glucose concentrations, they are subject to a physiological lag relative to blood glucose. During periods of rapid glycemic fluctuation, including instances of surgical stress, insulin administration, or hemodynamic instability, this lag may become clinically significant.^26^ Furthermore, vasoconstriction induced by anesthetic agents or vasoactive drugs may impair subcutaneous perfusion, thereby reducing sensor accuracy.^27^ Hypothermia, particularly during cardiopulmonary bypass, further exacerbates these effects by altering enzymatic sensor kinetics and tissue glucose diffusion.^28^ These mechanisms collectively explain why CGM performs optimally under stable physiological conditions but exhibits reduced reliability during extreme intraoperative perturbations.

The clinical implications of these findings are substantial. In contemporary anesthesiology practice, intraoperative glucose monitoring remains largely dependent on intermittent sampling, which inherently risks missing transient but clinically significant glycemic excursions. Our results demonstrate that CGM can detect a substantial proportion of hypoglycemic events that would otherwise remain unrecognized. Given the well-established association between hypoglycemia and adverse outcomes, particularly neurological injury and increased mortality, this represents a meaningful advancement in perioperative patient safety.^29^ Moreover, the observed improvement in TIR suggests that CGM may facilitate tighter and more physiologically appropriate glycemic control, potentially reducing glycemic variability. This factor is significant as variability itself has been linked to worse outcomes in surgical and critically ill populations.^30^

From a practical standpoint, integration of CGM into intraoperative workflows could enable continuous, data-driven insulin titration strategies, moving beyond the reactive paradigm imposed by intermittent monitoring. Devices such as Dexcom G6^®^ and Libre 2^®^ offer real-time trend information and alarms, which may assist anesthesiologists in anticipating glycemic excursions rather than merely responding to them. This capability is particularly relevant in prolonged or high-risk surgeries, where metabolic instability is common. However, given the observed limitations in accuracy under certain conditions, CGM should currently be considered adjunctive rather than replacement technology, requiring confirmation with venous or capillary measurements when clinical decisions carry high risk.

## LIMITATIONS

This systematic review carries methodological constraints that must be interpreted within the context of the available evidence. The aggregate sample across included studies remains numerically limited, and the restricted number of RCTs narrows the inferential capacity of the meta-analytic findings, particularly regarding causal relationships between CGM use and clinical outcomes.

Considerable between-study heterogeneity was identified throughout the quantitative synthesis, attributable to meaningful variation in patient case mix, device platforms, sensor calibration approaches, and perioperative glycemic management protocols. Such variability introduces uncertainty into pooled estimates and limits the generalizability of findings across distinct surgical contexts and institutional environments.

The absence of blinding in the majority of included studies represents an additional source of concern. Without allocation concealment from care providers and outcome assessors, both measurement bias and performance bias remain plausible, potentially inflating the apparent benefits associated with continuous monitoring in the intervention arms.

Reporting quality across studies was also inconsistent with respect to sensor-related technical events. Incomplete documentation of signal interruptions, calibration failures, and device malfunctions likely underrepresents the practical challenges inherent to CGM deployment in the operating room, which may differ substantially from ambulatory or outpatient settings where these sensors were originally validated.

Perhaps most critically, none of the eligible studies were powered to evaluate hard clinical endpoints. The current evidence base does not permit definitive conclusions regarding whether CGM-guided intraoperative glycemic management translates into measurable reductions in surgical morbidity or other patient-centered outcomes.

## CONCLUSION

The results of this meta-analysis support the view that CGM represents a meaningful advancement in intraoperative glycemic surveillance, offering adult surgical patients a level of metabolic oversight that intermittent POC measurement is structurally incapable of providing. The most clinically consequential contribution of CGM in this context lies in its capacity to identify hypoglycemic episodes that would otherwise go undetected throughout the course of anesthesia, alongside a demonstrated improvement in the proportion of operative time spent within the target glycemic range.

It is equally important to acknowledge that the analytical performance of these devices is not uniform across all surgical scenarios. Physiological conditions associated with profound hemodynamic and thermoregulatory disruption, as observed in cardiac procedures requiring hypothermic extracorporeal circulation, substantially attenuate sensor accuracy and impose meaningful constraints on clinical reliance upon CGM-derived readings in those specific settings.

The evidence bases currently available, while encouraging, reflects studies of limited scale and methodological heterogeneity. Whether the glycemic benefits documented in this review ultimately translate into reductions in postoperative morbidity or other patient-centered clinical endpoints cannot yet be determined from the existing literature. Addressing this question will require prospective, adequately powered trials designed specifically for the anesthesiology context, with sufficient sample sizes to detect clinically meaningful differences in surgical outcomes. Concurrently, the field would benefit from the development of standardized protocols governing sensor placement, calibration procedures, and alarm thresholds adapted to the intraoperative environment, so that future implementation efforts rest on a coherent and reproducible methodological foundation.

## Data Availability

All data produced in the present work are contained in the manuscript

